# AAEDM: Theoretical Dynamic Epidemic Diffusion Model and Covid-19 Korea Pandemic Cases

**DOI:** 10.1101/2020.03.17.20037838

**Authors:** Amang (Song-Kyoo) Kim

## Abstract

This paper deals with an advanced analytical epidemic diffusion model which is capable to predict the status of epidemic impacts. This newly propose model well describes an epidemic growth and it could be widely applied into various topics including pathology, epidemiology, business and data sciences. The Advanced Analytical Epidemic Diffusion Model (AAEDM) is a dynamic diffusion prediction model which is theoretically intuitive and its tractable closed formula could be easily adapted into versatile Bigdata driven analytics including the machine learning system. This dynamic model is still an analytical model but the periods of prediction are segmented for adapting the values from the dataset when the data is available. The epidemiologically vital parameters which effect on the AAEDM are also introduced in this paper. The evaluation of this theoretical model based on the Covid-19 data in Korea has been accomplished with relative fair future prediction accuracies. Although this analytical model has been designed from a basic exponential growth model, the performance of the AAEDM is competitive with other Bigdata based simulation models. Since the AAEDM is relatively simple and handy, anyone can use this model into analyzing outbreak situations in his daily life.

## 1. Introduction

As of March 12th, 2020, the WHO (World Health Organization) announced that the Coronavirus (aka. Covid-19) was deeply confirmed as a pandemic [1] which is the 3rd announcement of the pandemic by WHO since 1948 [2]. A pandemic is a disease that is spreading in multiple countries around the world at the same time [1]. The same news has been announced in several Korean newspapers and TVs [2]. On the next day, Korea had reached more than 8,000 persons are infected by the Corona virus [3]. It has been panicking in Korea and still Koreans have panicked. Politicians have started to appeal their opinions about the Covid-19 pandemic for the election on this year. News medias also have been involved to appeal their political opinions even based on scientifically fake information. Many professionals in the field of Epidemiology keep talking about the Covid-19 pandemic issues and strongly insisted their opinions to appeal their existence. Many people who do not have related knowledge could only trust what epidemiologists have said [4-5]. Although professionals have provided historical (scientific) facts which they might learn from the textbook or statistical data, there are a lot of conflictions even within their documented scientific facts. Many data scientists are working for delivering the valuable insights from the Covid-19 data [3, 6-7] but most of them are spending their efforts for gathering and visualizing data rather than doing theoretical studies to build up a model which could describe the Covid-19 pandemic situations. Basically, the area of Epidemiology is too complicated to provide solid solution based historical facts without having theoretical thinking at the beginning.

In the other hand, a virus epidemic is atypical textbook example of an exponential growth [9]. An exponential growth model insanely fits well in diffusion (or growth) problems including virus generating systems if the proper parameters are pre-defined at the beginning. But the main problem is that these parameters keeps changing in real-world problems like Covid-19 epidemic cases. The AAEDM (Advanced Analytical Epidemic Diffusion Model) is a dynamic diffusion prediction model which is still a theoretical model but the periods of prediction are segmented for adapting the values from the dataset when the data is available. Additionally, two critical parameters are suggested to build this new prediction model. These two parameters have been opted by most of data scientists and epidemiologists. These factors become vital parameters which could impact on the future prediction of a epidemic and give the clear cut to understand the current pandemic situation well.

This article consists of six sections. Section 2 describes the AAEDM which is an enhanced mathematical epidemic diffusion model to predict the number of infected people by typical epidemics. It also introduces the *kappa* and *zeta factors* which are capable to determine the characteristics of this prediction model. These factors could be used for understanding the current Covid-19 pandemic situations without analyzing heavy amount of dataset which only available for related professionals. Section 3 provides for data scientists or data analysts who want adapt the AAEDM into their statistical systems or the machine learning systems. There are couple of examples to simplify for gathering data and they could freely modify based on their circumstances with only one solid condition. Section 4 discusses the testing and validation of the AAEDM by using the dataset from the Covid-19 statistics in Korea. Although the AAEDM is a simple and handy model, the performance (mainly, an accuracy of future predictions) is fairly acceptable to be used for a statistical model by using Bigdata. Finally, the conclusion and further discussion regarding this research are provided in Section 5 and 6.

## 2. Enhanced Mathmatical Epidemic Diffusion Model

A conventional mathematical diffusion model is based on an exponential growth function which has been widely used on the fluid mechanics and thermal dynamic physics. The growth of virus epidemic is a textbook example of this kind of growth [9] and atypical expression is as follows [9]:

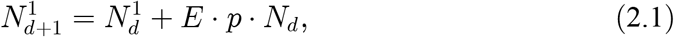

where

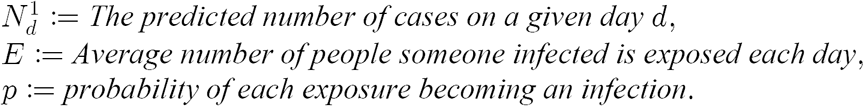

Acceding to various Algebra textbook sources, a prediction model of the exponential growth is as follows:

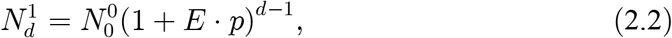

where 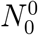 is the actual number of cases on a starting day from the real dataset. Although this formula insanely fits well in an epidemic case but the main problem of (2.2) is that some fixed values (*E* and *p*) keep changing in time to time even these values are supposed to fixed at the beginning of a growth prediction model.

### 2.1. Describing the present time based on an epidemic diffusion model

This paper suggests an enhanced prediction model of virus infections and the general growth model is described as follows:

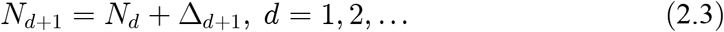

where Δ_*d*_ is typically known as the difference between two growth numbers (i.e., | *N*_*d*_ − *N*_*d−1*_ |) but this paper is newly proposing Δ_*d*_ as the function with some intersting parameters which have been usually ignored by epidemiologists. The formula of the newly proposed predicted value 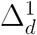 is as follows:

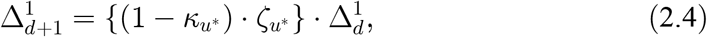

and the parameter 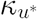 (i.e., *k-factor*) that presents the ratio of cured people within the net increased people could be found as follows:

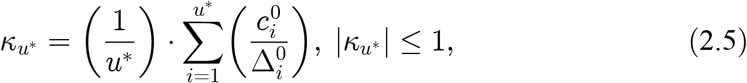

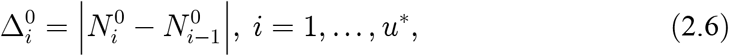

where

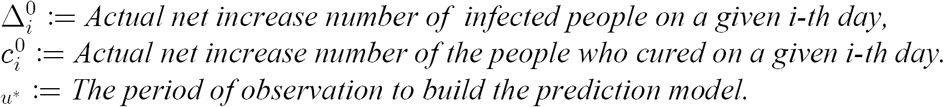

The parameter 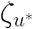 (i.e., ζ*-factor*) is the ratio of net increasing between the present and the past. The formula of this parameter is as follows:

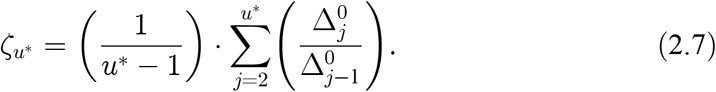

From (2.4)-(2.7), the predicted number of the net increase on a given day as follows:

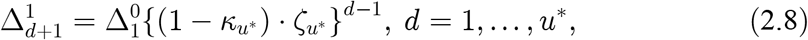

from (2.3) and (2.8),

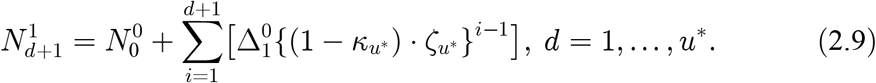

Finally, we have

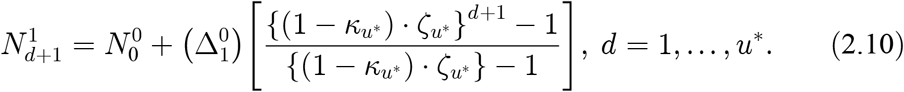

### 2.3. Kappa and Zeta: Cure and Net Increase Factors

This paper proposed two important factors which determine the characteristics of the AAEDM based prediction. The first parameter is the cure factor (aka. *kappa-factor, k-factor*) which is already shown from (2.5). The kappa-factor is an (daily) increase ratio between the net increase of cured people and the net increase of infected people (see Figure 1).

**Figure 1.**
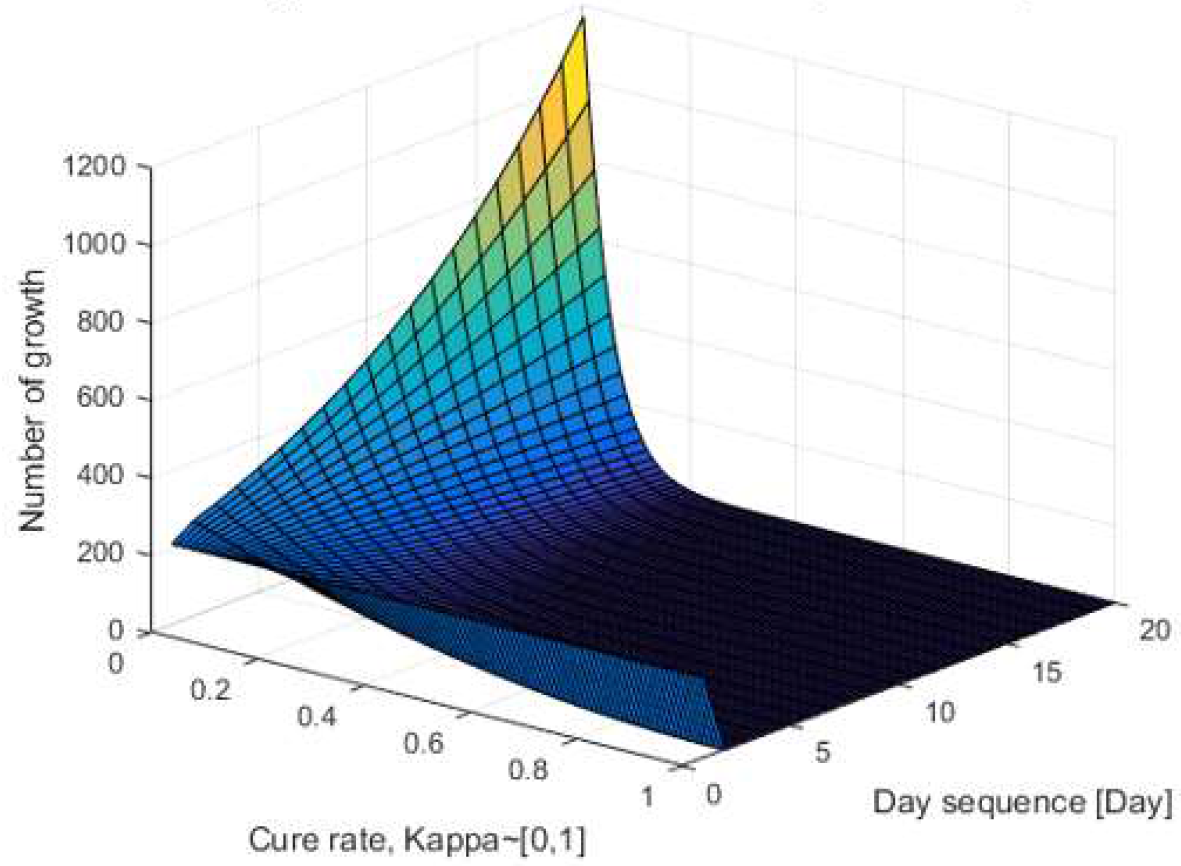
3D graph for the cure factor 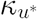.

From (2.5) and (2.7)-(2.8), the factor impacts on the total number of infected cases. Based on an empirical hunch, the critical threshold of the *kappa-factor* is 0.5 which indicates the speed of curing is more than a half of net epidemic increasing. As shown in Figure 2, the net increasing of spreading viruses is dramatically dropped when the kappa-factor is greater than 0.5 (i.e., 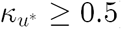) The current cure factor in Korea is around 0.09 as of March 14th, 2020.

**Figure 2.**
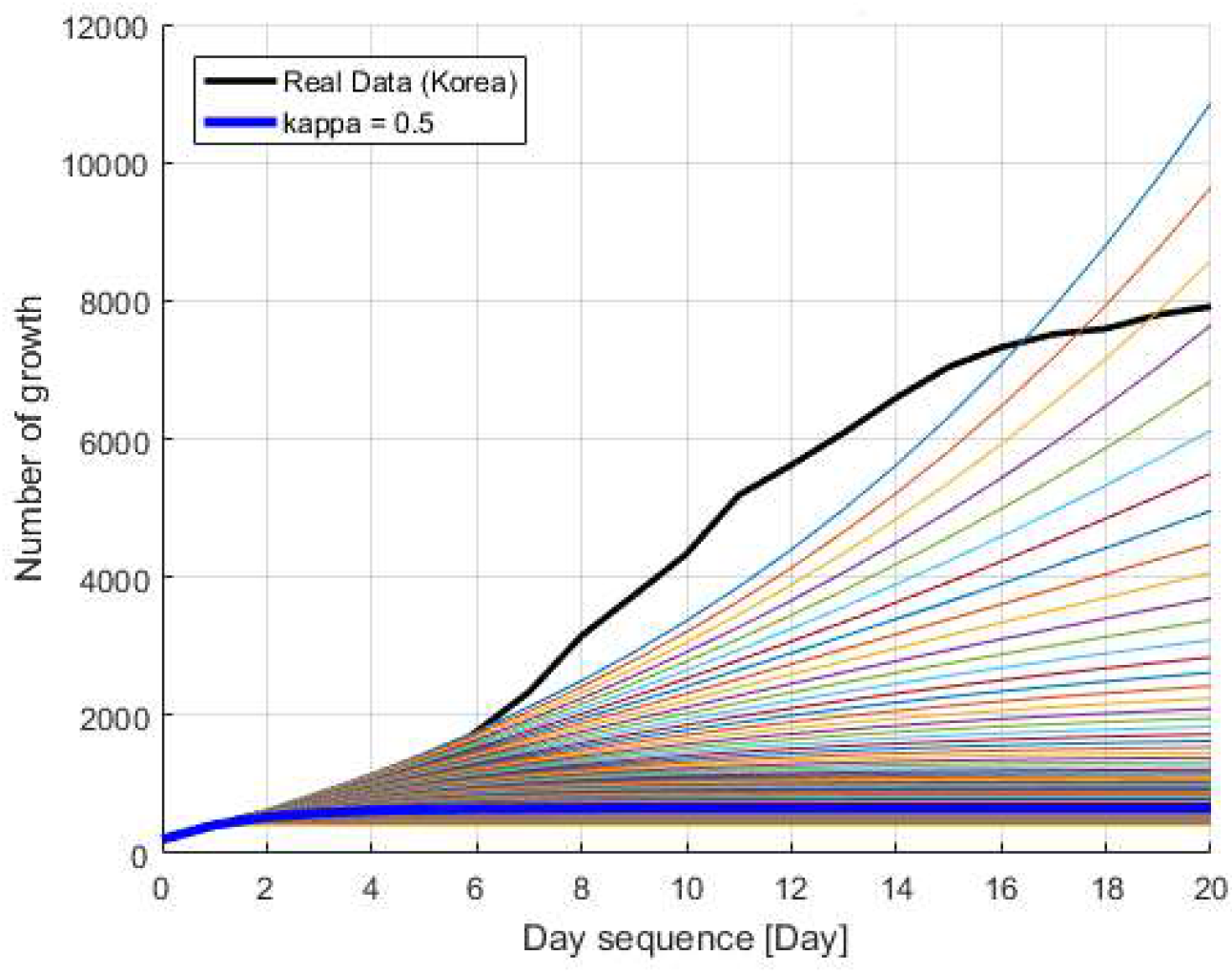
2D graph to show the impact of 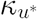.

Another critical parameter of the AAEDM is the net increase ratio factor which is called as *zeta-factor* (ζ*-factor*). The *zeta-factor* indicates an average value of the increasing ratio between the present and the past (see Figure 3).

**Figure 3.**
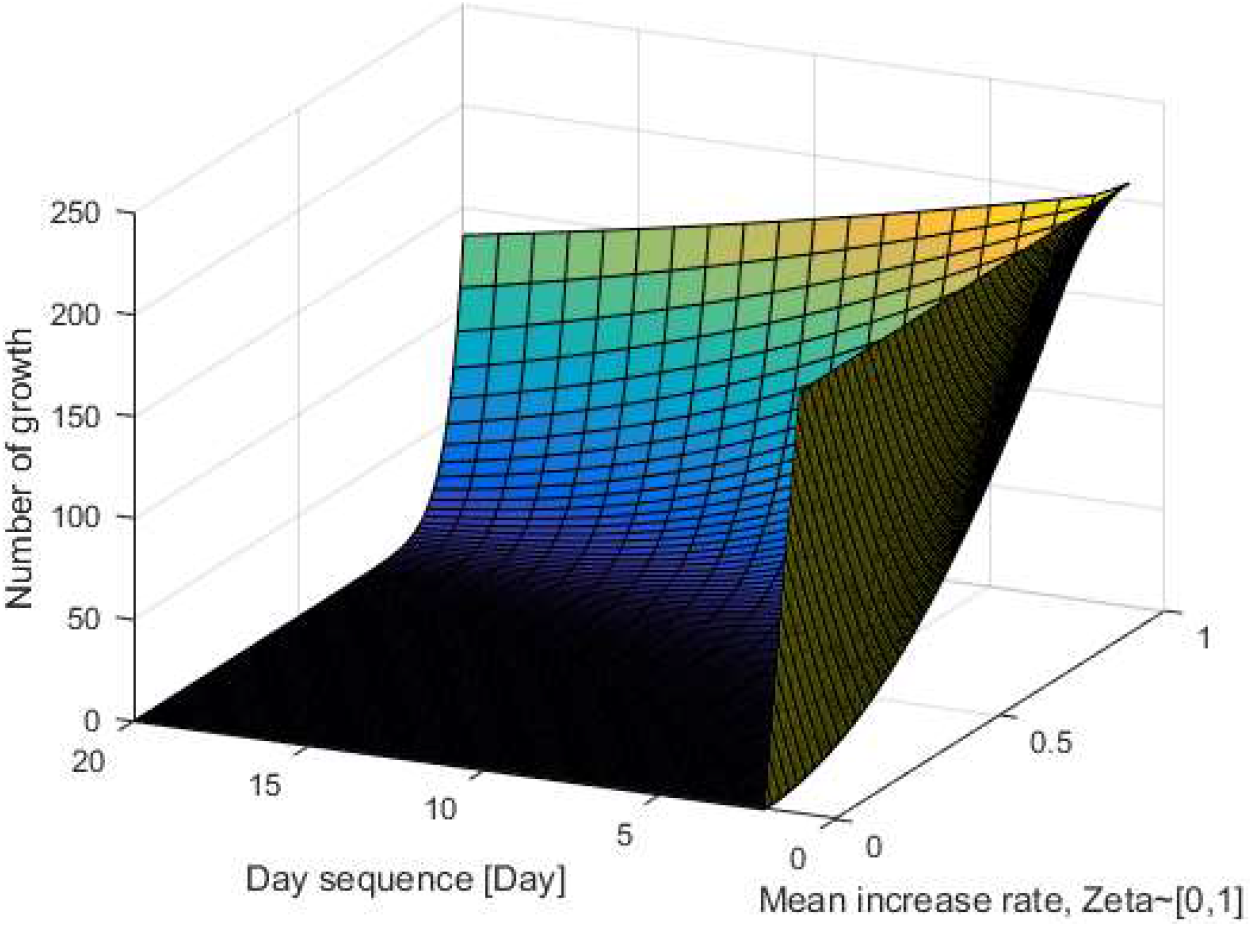
3D graph for the net increasing factor 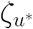.

The ζ*-factor* 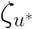 gives a good indication of a current status of epidemic growth. 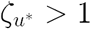 means the epidemic will be exponentially increased during a next prediction period (*d*^1^ ∈ [*u*\*, 2*u*\*]). In the other hand, 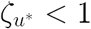 means that the growth will start saturating in the next prediction period. It is noted that the number of growth still will be increased even in the case of 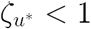. If 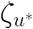 is closed to 1 (i. e., 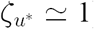), the growth tends to be linear.

### 2.3. Developing the future prediction model

Let us assume that 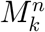 is the predicted accummulated number of infected people on a given day *k* ∈ [0, *u\**] in the prediction cycle *n*. The typical value of *n* is 1 which means that the collected dataset has been applied just before a future prediction. The prediction model for the next cycle (i.e., *n =* 1) could be dveloped from (2.10) as follows:

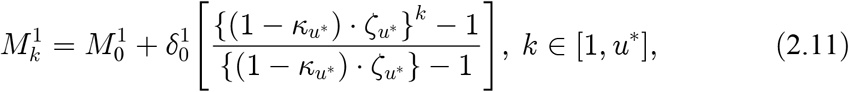

where

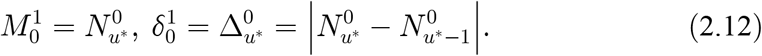

The actual data of the end of observation (or training, analysis) period becomes an initial value of a prediction model. The AAEDM is designed to adjust the initial conditions based on the recent updated actual data in every observation cycle whenever actual data are available during a previous prediction period:

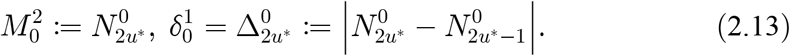

Hence, the model tends to be really accurate when the observation cycle is minimum (i.e., *u*\* = 1) and this special model will be treated in Section 4.1. Similarly, the predicted number of net-increasing of infected people could be determined as follows:

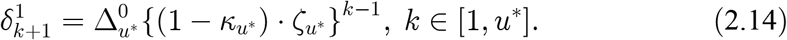

## 3. Parameter Approximations for Statistical Learning

Although many researchers are dealing with huge amount dataset and bring nice and fancy visualization based on these data [10-11] but the dataset of the *kappa-factor* and the *zeta-factor* may not fully available because these factors have been lightly dealt by data scientist. If users want to use this new theoretical prediction model, these parameters should be gathered from originally available dataset. This section provides the certain methods to simplify these two parameters.

### 3.1. Approximation of the parameters

The AAEDM has been designed for statistical analysis under extremely limited available datasets. Therefore, these two parameters (*k-factor* & ζ*-factor*) could be alternative obtained even the values are not calculated from the original formula. There are a lot of alternative approximation approaches regarding these parameters and one of typical approaches are as follows:

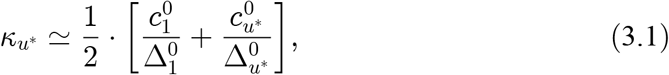

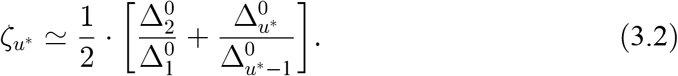

These parameters could be used for prediction models to estimate the number of accumulated infected people (2.10) and the net increasing (2.13). It is noted that these parameters are flexible enough to be modified based on data availability. Dataset might contain only recent values for these two parameters which might be missed at the beginning (e. g., missing of 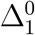 and 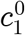). The values could be determined in different calculations such as:

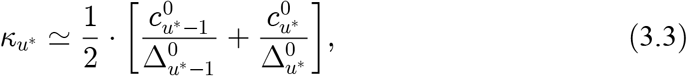

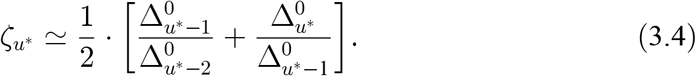

The way of modifying these two factors depends on various conditions from researchers. As long as the rationales of these parameters are same, researchers could modify these parameters as much as they want. The *kappa-factor* and *zeta-factor* could be even more simplified as follows:

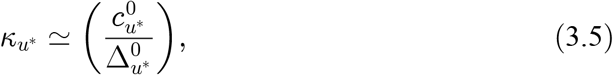

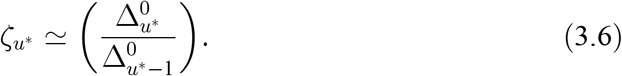

The method of calculating these two factors could be mixed and matched. For instant, a data analyst could determine 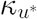 from (3.1) and 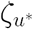 from (3.4) instead of using the original forms from (2.5)-(2.6). Other scientists might differently calculate these factors from (3.1) and (2.7). The optimal values of these factors could be determined by currently available datasets. The methods for determining these factors are freely chosen except for one condition: *Consistencies*. It means that the formula for calculating *k* and ζ factors should be the same for both observing (or training) presents and predicting futures. If the factors are calculated in a certain way at the beginning, the same way of calculation should be applied into predicting the future. The data analysis for the proof of the AAEDM chooses the *k*-factor from (3.5) and ζ-factor from (2.7).

## 4. Korea COVID-19 Pandemic Case Studies

The dataset for Korea case has been collected from various sources [3, 6-8] for developing the statistical testing but all data have used the same original source from the Korean government [8]. The data have been collected as daily basis since February 21, 2020 until March 14th, 2020. Since, the data are scattered as daily reporting documents, the actual values are manually collected. Fortunately, it does not require heavy amount of this data because the purpose of data collection is for the AAEDM demonstration. It is noted that the data collection from the government is confused because they change the way of delivering numbers. During the data collection period, they started counting the total number of infected people without adding the number of cured people. The total number of infected people had been counted including the number of cured in the past. The values in the dataset has been revised by adding the number of cured people after certain period of time.

### 4.1. Minimum observation period

Since the model deals with the ratio, it requires to observe at least two measure from the actual dataset. From (2.10) and (2.12)-(2.13), the AAEDM is simplified as follows:

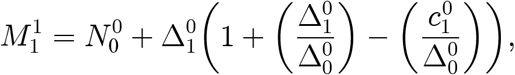

where

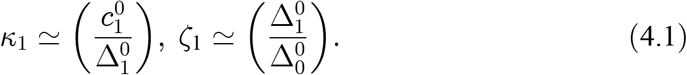

As shown in Figure 4, the results are fairly accurate (see Figure 4) because the initial values for prediction have been measured everyday but it could predict only one day after. It is noted that this special AAEDM is capable to predicts an accurate future with only three values which everyone could find exact estimation even from news media [3, 6-7] or from the government reports [8]. The accuracy of this AADEM is more than 90 % (≃ 0.9175).

**Figure 4.**
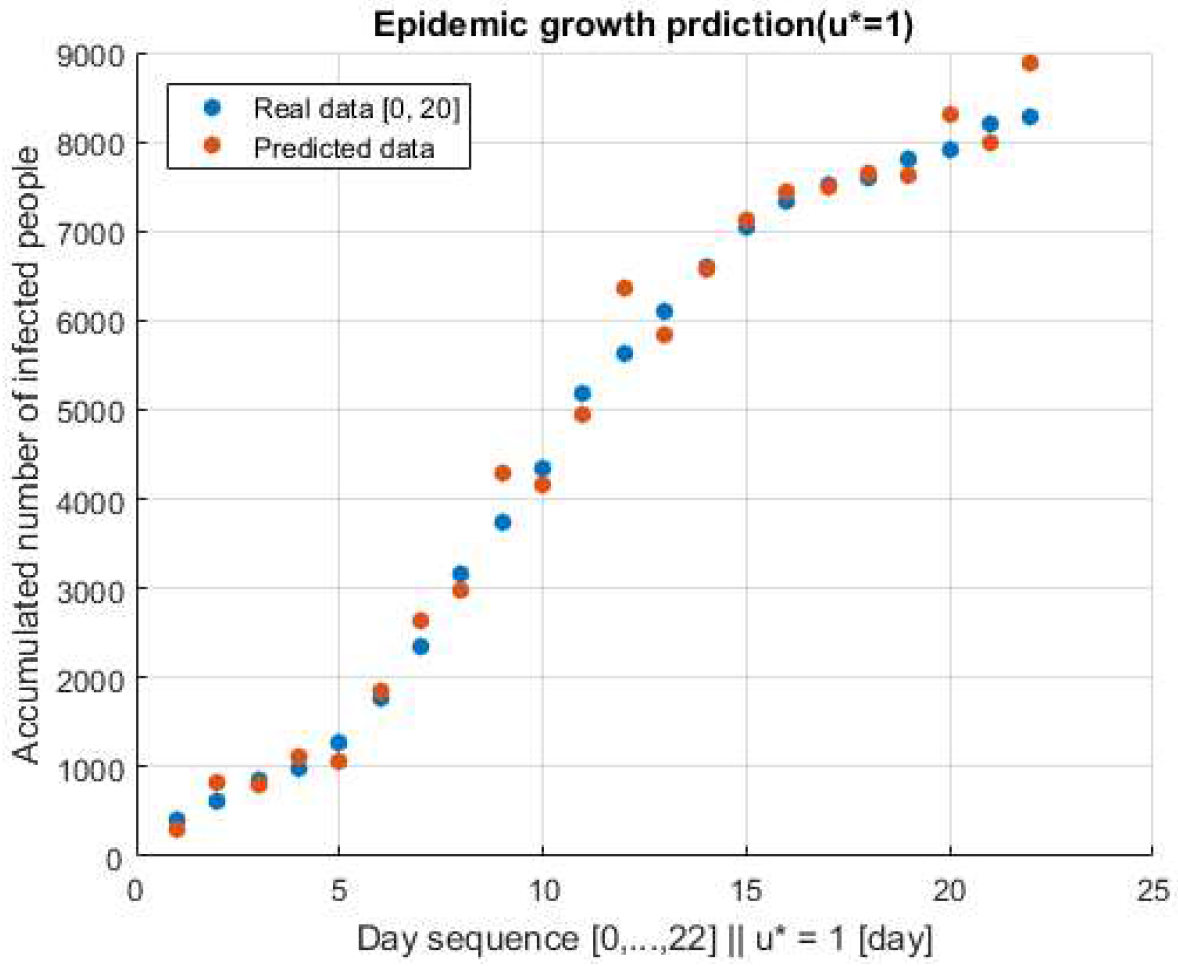
The special AAEDM [2020.02.21-03.14].

### 4.2. The 10 days observation and the next 10 days predictions

The calculation by using the AAEDM is straight forward and the predicted value could be estimated from (2.10). The 10 days (actually 11; 0,1, …, 10) during the observation period (02.21-03.02) is selected for the observation (i.e., a training phase in a machine learning system). All important parameters including the *k* and ζ could be calculated by using the data during this period.

The comparison result is shown in Figure 5 and the accuracy of the theoretical model is 0.8676 (≃ 87 %) which is not that bad to get the expected values directly from theoretical formula. The future prediction starts at the end of the observation period and the parameters are adjusted based on the real data. The period of the prediction is the next 10 days from March 2nd to 12th. The predicted values have been compared with the actual data after passing 10 days and the accuracy is around 83% (see Figure 6).

**Figure 5.**
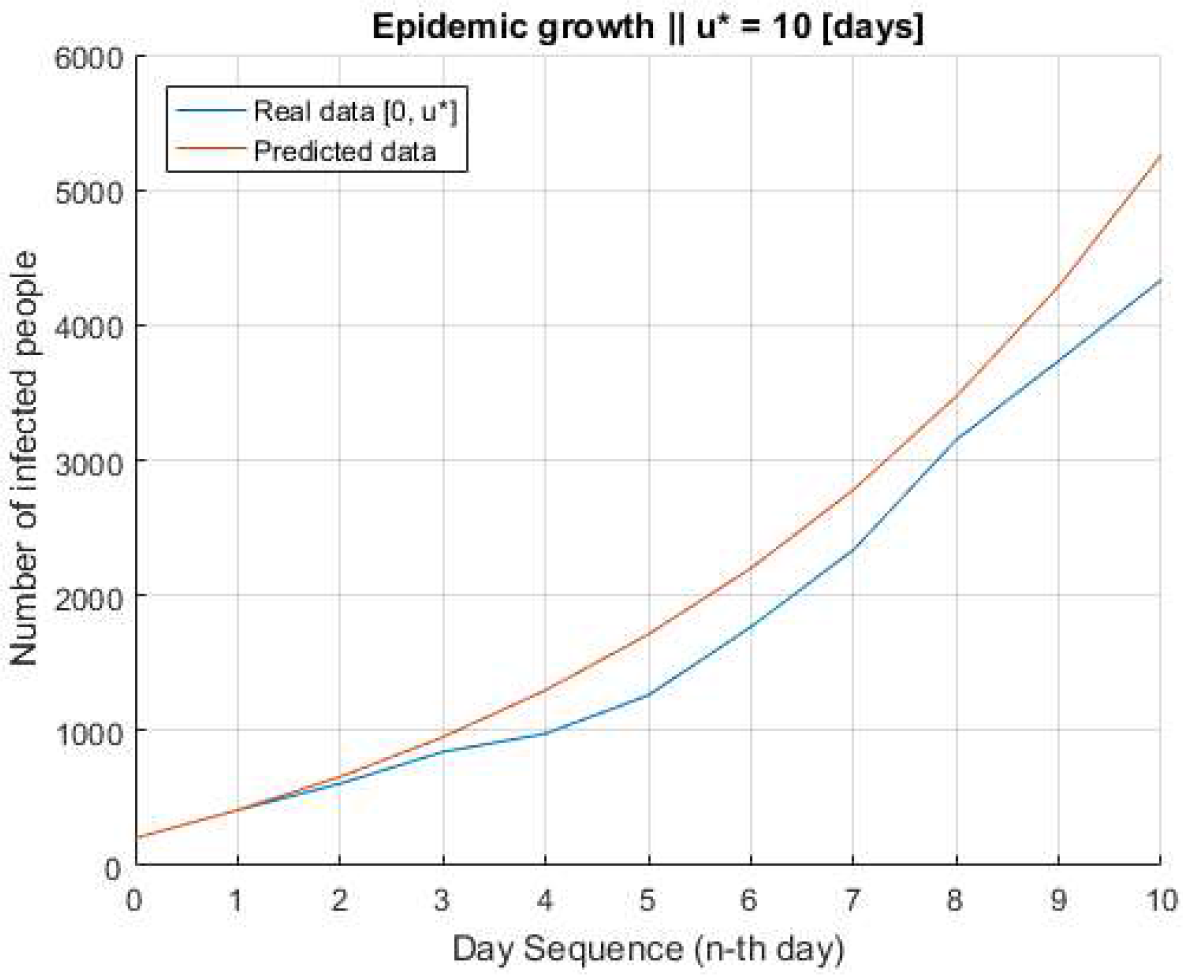
Comparison result with the real data during the observation.

**Figure 6.**
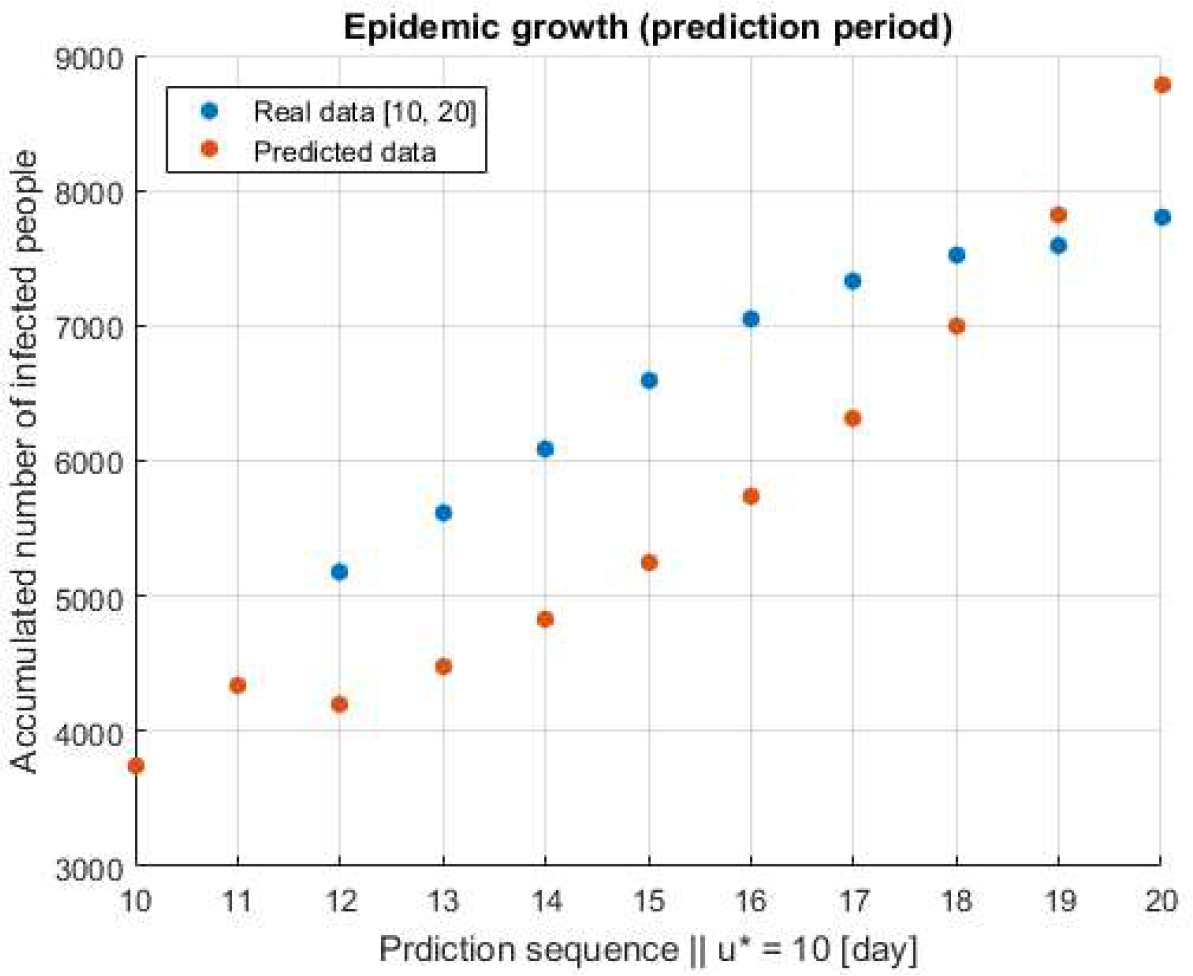
Prediction result with the next 10 days.

## 5. Conclusions

Korea is still under suffering of the Covid-19 pandemic and many peoples who live in Korea are still on panic. Many news and social medias have produced the related news and comments based on unscientific facts. Even documented scientific facts bring a lot of confusion because of conflicted facts by their own. The purpose of this paper is removing these confusions by understanding the basics of nature. The way of understanding the Covid-19 epidemic status is just checking two factors in daily basis. It will give the fairly accurate status of the epidemic. As of today, the value of *kappa-factor* is around 0.08 and the value of *zeta-factor* is 1.14 and the accumulated number of infected people might be increased up to 12,000 at the end of March. News and social medias should not be fully trusted because they will say whatever they want people to believe. The world including Korea will be eventually settled and safe from the Covid-19 pandemic when the *kappa-factor* is greater than 0.5 and the *zeta-factor* is lower than 1 (i.e., 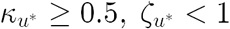). That is what the AAEDM talks today.

## 6. Future Directions

This section is the list of future research which researchers might be interested into adapting or initiating their research projects.

### Adapting the AAEDM into other countries and other regions

Although the AADEM has been adapted for proofing the concept in Korea case, it could be adapted into other countries or other regions such as Italy, USA, UK, UAE and so on. They will understand the Covid-19 situation more clearly by tracking the *kappa* and the *zeta* factors.

### Improving the AAEDM

The closed formula of the AAEDM is still in the basic stage. Other vital factors such as social distancing [12] could be added and evaluated with real epidemic data.

### Optimizing the critical parameters in the AAEDM

Researchers could find the critical parameters in the AAEDM including *k*, ζ and *u*^*\**^ by using the machine learning (ML) techniques.

### Finding the analytical function of *k* (*t*) and ζ (*t*)

In the AAEDM, the *k* and ζ factors are determined as a constant value most based on empirical hunch. But the author strongly believes that *k* (*t*) and ζ (*t*) are time dependent stochastic functions. It could not be easily found analytically but the ML technique could build up the functions based on massive data analytics.

### Finding the impacts of kappa and zeta factors

Although these two parameters are epidemiologically important and key elements of the AAEDM formula, these factors are coming to the picture by empirical hunch and it is not statistically proven yet. Researchers might be interested in finding statistical relationships (i.e., correlation, p-value, hypothesis testing and so on) between the epidemic growth and these two parameters based on massive amount of data analytics.

### Experiment practices for SETM kids

Practicing scientific experiments is one of important activities for STEM kids. As it might be noticed, the basic model of the AAEDM is simple and handy. Everyone who knows about an exponential growth can easily understand the AAEDM for their scientific experiments.

All topics above are only small parts of the potential research topics based on the AAEDM and researchers may feel free to use the AAEDM for their own purposes as long as this paper is properly citied.

## Data Availability

All data is publicly available.

https://github.com/amangkim/aaedm-covid19-kr

## Acknowledgment

The source codes and the dataset for proofing the concept are available on the GitHub** and anyone can freely use for their own purposes but only with the proper citation of this paper. It is also noted that the values in the dataset might not be fully trusted because of manual data correction.

## Diclaimer

The paper is preliminary report of work that have not been peer-reviewed. It should not be relied on to guide clinical practice or health-related behavior and should not be reported in news media as established information.

## Footnotes

** (Available on April, 2020) https://github.com/amangkim/aaedm-covid19-kr

